# Double burden: fatigue and poor sleep quality comorbidity and its predictors among cancer patient in Amhara region, northwest Ethiopia: institutional based cross-sectional study design

**DOI:** 10.1101/2025.11.13.25340178

**Authors:** Gebreeyesus Abera Zeleke, Astewil Moges Bazezew, Birtukan Atena Negash, Desalegn Getachew Ayele, Yalemwork Getahun Azanaw, Alamirew Enyew Belay, Alebachew Ferede Zegeye

## Abstract

**Background:** Cancer-related fatigue and poor sleep quality are among the most prevalent and distressing symptoms experienced by patients with cancer, significantly impairing physical, emotional, and cognitive functioning. Despite their high prevalence and detrimental impact on quality of life, the comorbidity of fatigue and sleep disturbances remains underexplored, particularly in low-resource settings where access to comprehensive oncology care is limited. Understanding the magnitude, contributing factors, and interrelationship of these symptoms is essential for developing targeted interventions. However, existing research predominantly focuses on either fatigue or sleep quality in isolation, highlighting a critical gap in evidence regarding their concurrent occurrence and synergistic effects on cancer patient.

**Method:** Institutional-based quantitative cross-sectional study was conducted among adult cancer patients receiving cancer treatment at an oncology unit from May to June 2025. A systematic random sampling technique was used to select 422 samples. After obtaining consent data were collected using a structured Interviewer-administered questionnaire. Then data were entered into Epi-data version 4.6 and exported to Stata version 14 for analysis. Model fitness was checked by the Hosmer-Lemeshow goodness of fit test. Descriptive statistics including, frequencies and proportions were computed and presented by using tables and texts. Bivariable and multivariable logistic regression analysis was computed considering p<0.05 to be statistically significant at the final model.

**Result:** A total of 405 cancer patients were included in this study, of whom 46.67% experienced comorbid fatigue and poor sleep quality. Age 61-89 years [**AOR** = 2.79, 95% CI: [1.02, 7.62]. Rural residency [**AOR** = 2.03 95%, CI: [1.02, 4.01], Married & divorced [**AOR**= 2.65 95%, CI: (1.01, 6.90)] and [**AOR**= 3.54 95% CI: (1.10, 11.40)], Inpatient [**AOR**=2.84, 95%, CI: (1.63, 4.95)]. Stage II and Stage IV [**AOR**=3.92, 95%, CI: ([1.89, 8.12] and [AOR= 2.52, 95% CI: (1.04, 6.15)] respectively, cancer duration [**AOR**=2.70, 95% CI: (1.14, 6.39)]. Anxiety [**AOR**= 1.93, 95% CI: (1.06, 3.51)]. depression [**AOR**= 2.10, 95% CI: (1.19, 3.70)].

**Conclusion and recommendation:** Comorbidity of fatigue and poor sleep affected nearly half of cancer patients, representing a substantial and underrecognized clinical burden that necessitates systematic assessment and integrated, multidisciplinary interventions in oncology care. Age, Residence, Marital Status, Cancer Stage, Cancer Duration, Inpatient Admission, And Anxiety and Depression were significant predictors. These findings highlight the need for routine screening and integrated interventions targeting both physical and psychosocial determinants, alongside strengthening supportive and multidisciplinary care to improve patient outcomes.

## Introduction

The comorbidity of cancer-related fatigue (CRF) and poor sleep quality constitutes a critical and debilitating symptom cluster, defined by persistent, non-relieving exhaustion and disrupted sleep processes, which together amplify clinical complications, intensify functional impairment, and severely compromise the quality of life in cancer patients. Cancer is major public health challenge, with nearly 20 million new cases and 9.7 million cancer-related deaths reported globally in 2022[1]. Beyond the rising prevalence of cancer, fatigue and sleep disturbances persist as among the most prevalent and distressing symptom clusters, representing a major source of symptom burden and significantly impairing patients’ quality of life[2, 3]. Research from the U.S. Centers for Disease Control and Prevention (CDC) and the World Health Organization (WHO) shows how these symptoms impair psychological health, physical functioning, treatment compliance, and general quality of life.[4, 5]. Longitudinal and systematic reviews repeatedly show that fatigue and poor sleep quality are not only highly prevalent but also commonly co-occur, exacerbating each other’s symptoms and making patients’ suffering worse[6, 7]. Furthermore, meta-analyses show that, depending on the disease stage and treatment approach, between 40 and 80 percent of patients experience cancer-related fatigue (CRF), and 50 to 70 percent experience clinically significant sleep disturbance [5, 8]. When these conditions co-occur, patients face synergistic declines in quality of life, reduced functional status, and poorer survival outcomes[9]. Additionally, longitudinal data demonstrates that untreated poorer and fatigue can last for years after treatment, severely reducing the chances of surviving. The intensity of this comorbidity has been linked to psychological morbidity, such as anxiety and depression, lower productivity, and higher healthcare consumption. [10]. Consequently, the combined burden of fatigue and poor sleep quality constitutes a significant but little-known aspect of cancer treatment worldwide. Numerous clinical, sociodemographic, and psychological factors have been found to predict the comorbidity of sleep disruption and cancer related fatigue. Higher risks of these symptoms are consistently linked to advanced cancer stage, longer time since diagnosis, and harsh treatment modalities like chemotherapy and radiation[11, 12]. Biological factors, including anemia, pain, systemic inflammation, and poor nutritional status, further exacerbate symptom severity[13]. Psychological factors, particularly depression and anxiety, are strong independent predictors, while low physical activity, poor social support, and rural residency have been implicated in low- and middle-income country (LMIC) settings[13, 14]. This complex etiology emphasizes the necessity of integrated, situation-specific management strategies. The comorbidity of cancer related fatigue and poor sleep quality is not well studied in sub-Saharan Africa, especially Ethiopia, despite its increasing international attention. The majority of Ethiopia’s current study has focused on these symptoms separately. Studies carried out in Ethiopia’s Amhara area and elsewhere, for instance, have found a high frequency of CRF and poor sleep quality independently, and correlations between these outcomes and variables like depression, advanced cancer stage, anemia, pain, and inpatient status have been found[15–17]. However, to our knowledge, no prior study in northwest Amhara has quantified the combined burden of fatigue and poor sleep quality as a binary comorbidity outcome. This represents a critical knowledge gap, as simultaneous evaluation provides more comprehensive insights into patient symptom clusters and may better inform integrated care strategies. In light of this evidence gap, there is an urgent need for studies that establish both the magnitude and determinants of fatigue–sleep comorbidity in Ethiopian oncology settings. Such evidence is particularly relevant to Ethiopia, where late-stage presentation, limited access to specialized care, and high psychosocial burden may amplify the prevalence and impact of these symptoms. Therefore, this study aimed to estimate the magnitude of comorbid fatigue and poor sleep quality and its predictors among adult cancer patients attending oncology unit in the northwest Amhara region, Ethiopia.

## Method and material

### Study design, and period

Institutional-based quantitative cross-sectional study was conducted among adult cancer patients receiving cancer treatment at an oncology unit and Data collection was take place over a month from May to June 2025.

### Study Area

The study was carried out in the cancer treatment hospitals in the Amhara region, Northwest Ethiopia. There are eight comprehensive hospitals in the in Amhara region, of which only four hospital have a cancer treatment oncology unit. Thus, hospitals were University of Gondar Comprehensive Specialized Hospital (UOGCSH), Felegehiwot Comprehensive Specialized Hospital (FCSH), Tibebegion Comprehensive Specialized Hospital (TCSH), and Dessie Comprehensive Specialized Hospital (DCSH). The distances of these hospitals from Addis Ababa, the capital city of Ethiopia, are 748 km, 564 km, 399 km, and 480 km, respectively. Each comprehensive specialized hospital serves 3.5–5 million people[18]. Each of these hospitals operates an oncology clinic or treatment center that provides both inpatient and outpatient services. Specifically, the oncology units of FCSH=28, UoGCSH=32, DCSH=20, and TCSH= 25 beds are equipped respectively. These services are delivered by a multidisciplinary team comprising nurses, oncologists, and general practitioners.

### Study Population

All adult cancer patients (aged ≥18 years) attending the oncology clinics during the data collection period was study population and study unit consisted of adult cancer patients who were chosen at random during the data collection period.

### Eligibility Criteria

#### Inclusion Criteria

Eligible participants were including adult patients (aged 18 years or older) with a confirmed diagnosis of any type of cancer. Patients must be either currently receiving active cancer treatment (such as chemotherapy, radiotherapy, immunotherapy, or surgery) or undergoing follow-up care. Additionally, participants must be willing and able to provide informed consent and demonstrate sufficient cognitive and communicative ability to complete the study questionnaire reliably.

#### Exclusion Criteria

Patients who are critically ill or unable to communicate, as well as those diagnosed with psychiatric disorders or cognitive impairments that interfere with reliable self-reporting, should be excluded. Additionally, patients with previously diagnosed primary sleep disorders that are not attributable to cancer or its treatment should be considered separately, as their sleep disturbances are independent of oncologic processes.

### Sample Size Determination

The sample size was calculated using the single population proportion formula, considering: Estimated prevalence (P) of comorbid poor sleep quality and fatigue among cancer patients 50% (due to lack of local data).

Prevalence (P)=50%

Confidence level = 95% (Z = 1.96). Margin of error (d) = 5% (0.05).

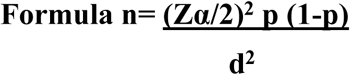

n=1.96*0.5(1-0.5)/ 0. 05^2^=384

Adding 10% non-response rate 422. Final sample size was 422.

### Sampling Technique

Systematic random sampling technique was used to select study participants from each comprehensive specialized hospital and proportional allocation for each hospital was properly calculated based on number of cancer patients they served per month. The sampling interval was determined by dividing the total study population who had follow-up and on treatment during one typical month (1500) by total sample size (422). Therefore, the sampling fraction was calculated to be 1500/422 ≈ 3. The first participant was selected randomly by a lottery method from 1-3 and the next respondent was chosen at regular intervals (every 3) by data collectors and patient register follow up log book, patient MRN No. and the patient itself should strictly use to avoid repeated data collection (**figure 1**)

**Fig 1:**
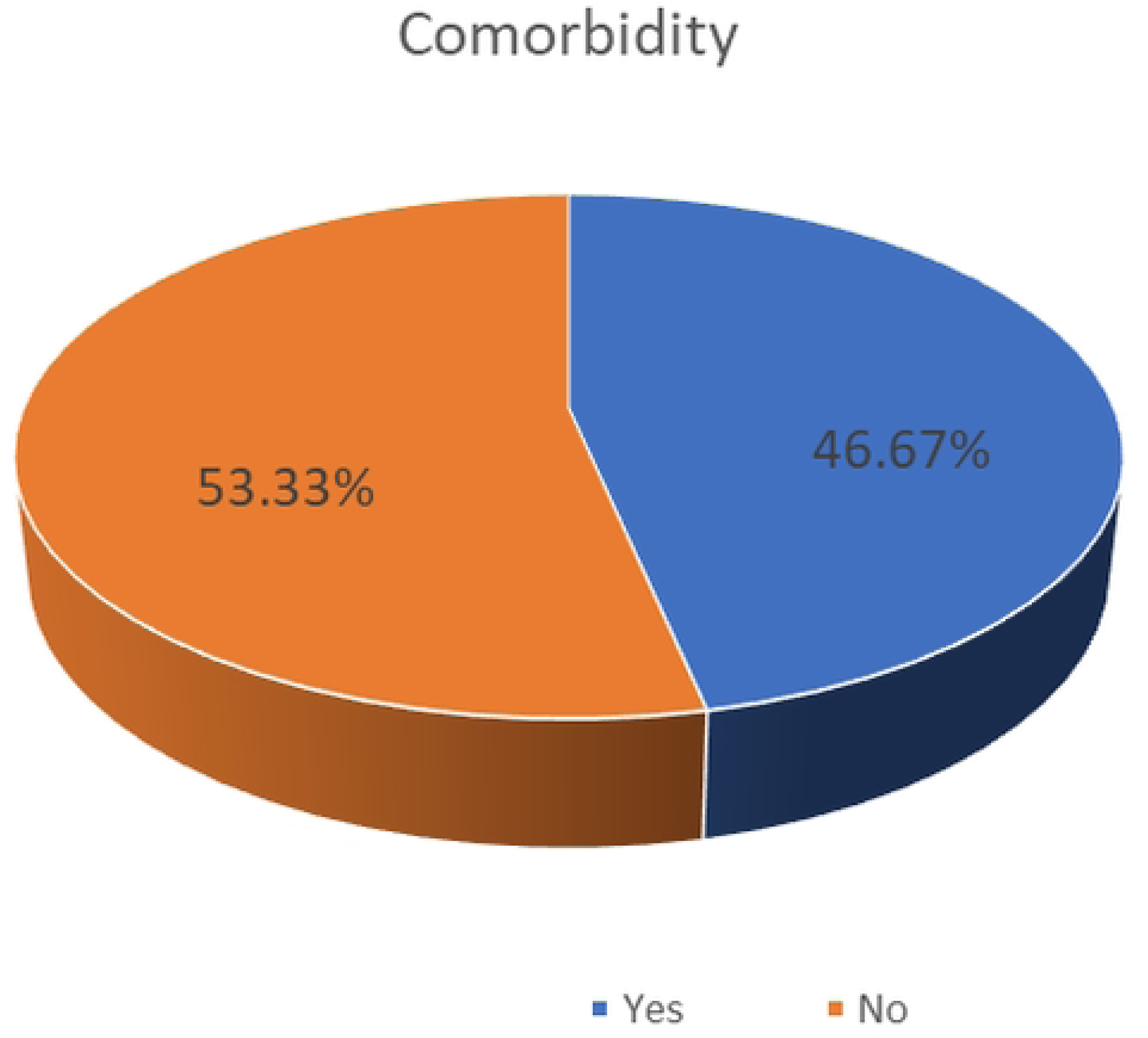
Schematic presentation of the sampling procedure comorbidity of fatigue and Poor sleep quality and its predictor among adult cancer patients, Northwest, Ethiopia, 2025

**FHCSH:** Felege Hiwot Comprehensive Specialized Hospital

**TGCSH:** Tibebe Gion Comprehensive Specialized Hospital

**DCSH:** Dessie Comprehensive Specialized Hospital

**UoGCSH:** University of Gondar Comprehensive Specialized Hospital

### Data Collection Tools and Procedures

Data were collected using a structured, interviewer-administered questionnaire with open-ended and closed-ended questions. There are eight parts to the data collection tool. Part one contains sociodemographic data, part two disease and treatment-related signs and symptoms, part three sleep quality assessment, part four Brief Fatigue Inventory. The tools for socio-demographic and clinical factors were adapted from the review of different pieces of literature. Anxiety and depression were assessed using the Hospital Anxiety and Depression Scale (HADS) [19] which was also validated in Ethiopian cancer patients [20]. Performance status was assessed by the single item Eastern Cooperative Oncology Group (ECOG) performance status scale [21]. Social support was assessed by the three-item Oslo social support scale (OSSS-3) [22]. Sleep quality was assessed by a standardized and validated Pittsburgh Sleep Quality Index (PSQI). The PSQI was designed to evaluate the subjective quality of sleep in the past month. It contains 18 self-rated questions, including seven subscale components (subjective sleep quality, sleep latency, sleep duration, habitual sleep efficiency, sleep disturbance, use of sleep medication, and daytime dysfunction). The global PSQI score ranges from 0 (no difficulty) to 21 (severe difficulties in all areas). Each component score ranges from 0 (no difficulty) to 3 (severe difficulty). Higher global and component scores indicate more severe complaints and a higher level of poor sleep quality. A global PSQI score greater than 5 yielded a diagnostic sensitivity of 89% and specificity of 86.5% (kappa = 0.75, P ≤ 0:001) in distinguishing “poor” from “good” sleepers [23]. The construct validity and internal consistency are further evaluated and supported in cancer patients with a Cronbach’s α value of 0.81 [24]. For present study, the internal consistency measurement of the PSQI subscales found a Cronbach’s alpha coefficient of 0.761 from the pretest data which was acceptable for this study. Brief Fatigue Inventory scale (BFI): Cancer patients who scored greater than or equal to four (≥ 4) moderate to severe in BFI measurement scale was considers fatigue whereas, < 4 in BFI scale was considers not fatigue[25–29].

### Data Collection Procedure

Data collectors (trained BSc nurses) conducted face-to-face interviews in a private setting to ensure confidentiality and comfort. Medical records were reviewed to obtain clinical data. The questionnaires were first translated into the local language (Amharic) and then back-translated into English to ensure accuracy. A pretest of the instruments was carried out on 5% of the sample at Debre Berihan Comprehensive Specialized Hospital in a similar population to identify and address potential issues

### Outcome variable

Outcome variable was a binary variable representing the co-occurrence (comorbidity) of poor sleep quality and cancer-related fatigue. Sleep quality was assessed using the Pittsburgh Sleep Quality Index (PSQI). The global PSQI score ranges from 0 to 21, with higher scores indicating poorer sleep quality. A commonly accepted cut-off score of >5 was used to classify poor sleep quality. Fatigue was measured using the Brief Fatigue Inventory (BFI). The (BFI) score ranges from 0 to 10. BFI score ≥ 4 were categorized as clinically significant fatigue, while those with scores < 4 were considered not clinically significant fatigued. Participants were classified as having the comorbid condition (coded as **1**) if they met both criteria simultaneously that is, they had a PSQI **score > 5** and a BFI score ≥ **4**. All other participants (i.e. those with either or neither condition) were coded as **0**, indicating absence of the comorbid outcome.

**Comorbidity of fatigue and poor sleep quality**: was defined as the presence of both clinically significant fatigue and poor sleep in the same individual. Fatigue was measured using Brief Fatigue Inventory, with a score of ≥ 4, indicating clinically significant fatigue. Sleep quality was measured using the Pittsburgh Sleep Quality Index (PSQI), with a global score > 5 indicating poor sleep. Participants meeting the criteria for both were coded as “1” (comorbidity); all others were coded as “0” (No comorbidity).

### Independent Variables

Sociodemographic such as Age, sex, residence, marital status, education, occupation. Clinical factors: Cancer type, cancer stage, duration since diagnosis, treatment modality, Behavioral and psychosocial factors: depression/anxiety.

### Operational Definition

**Comorbidity fatigue and poor sleep quality:** is defined as the simultaneous presence of poor sleep quality and fatigue[15]

**Good sleep quality**: a global PSQI score of ≤5[30]

**Poor sleep quality**: a global PSQI score of >5[30]

**Anxiety and depression:** A patient with more than 10 points on the Hospital Anxiety and Depression Scale (HADS) has anxiety and depression problem. **Good performance:** if Eastern Cooperative Oncology Group performance status which ranges from 0-4 (ECOG-PS). Patient score 0-1. **Poor performance:** if Eastern Cooperative Oncology Group performance status (ECOG-PS) patient score 2-4. **Social support**: by using the three-item Oslo social support scale (OSSS-3), a score of 3–8 represents ‘poor support’, 9–11 ‘moderate support’, and 12–14 ‘strong support[31, 32].

### Data Quality Assurance

Supervisors and data collectors received training on interviewing techniques, ethical considerations, and study objectives. To guarantee accuracy and consistency, completed surveys were supervised and reviewed every day. To reduce entry errors, data entry was done twice. Scales such as the PSQI and BFI were tested for internal consistency using Cronbach’s alpha.

### Data Processing and Analysis

Data were coded and entered into Epi Data and then exported to Stata version 14 for analysis. Descriptive statistics were used to summarize sociodemographic and clinical characteristics as well as the prevalence of comorbid fatigue and poor sleep quality. Bivariate analyses using Chi-square tests were conducted to assess associations between independent variables and comorbidity. Variables with a p-value < 0.25 in the bivariate analysis were included in the multivariable logistic regression model to identify independent predictors of comorbidity, and adjusted odds ratios (AORs) with 95% confidence intervals (CIs) were reported.

### Ethical Considerations

Ethical approval for this study was obtained from the Institutional Review Board (IRB) of the University of Gondar. Permission was also granted by the respective hospital administrations. Written informed consent was obtained from each participant after providing a clear explanation of the study’s purpose, potential risks, and anticipated benefits. To ensure confidentiality and anonymity, all data were securely stored and made accessible only to the research team.

## Result

### Socio-demographic characteristics of study participants

In this study, 405 patients were participated with 97% response rate. nearly two-thirds (60.74%) participants were female. The Mean age of participants were 46.43 with (Std. Dev. 15.43) years. Around one third of the participants (32%) were stage two cancer and in terms of residency, slightly more than half of the participants (62.96%) were from rural area and nearly half of the participants (45.43%) were not educated (**Table 1**).

**Table 1:**
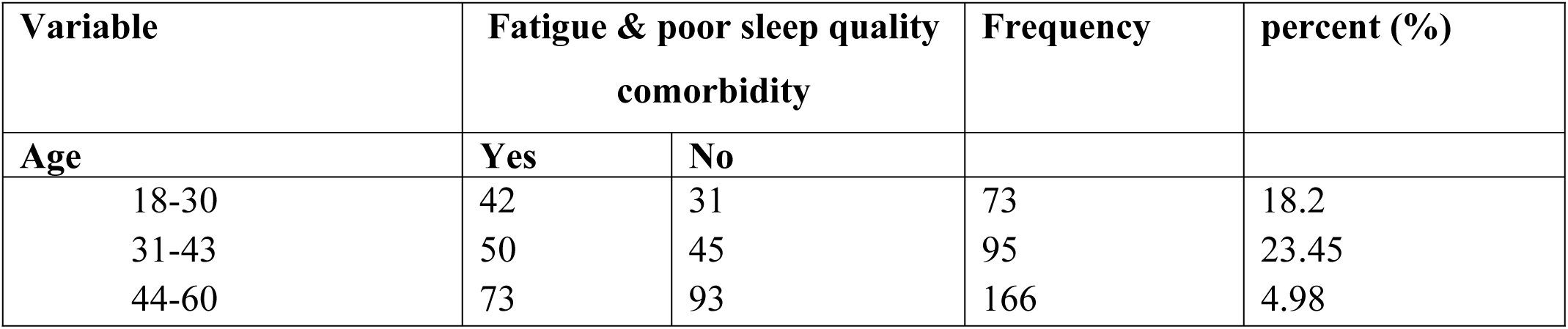
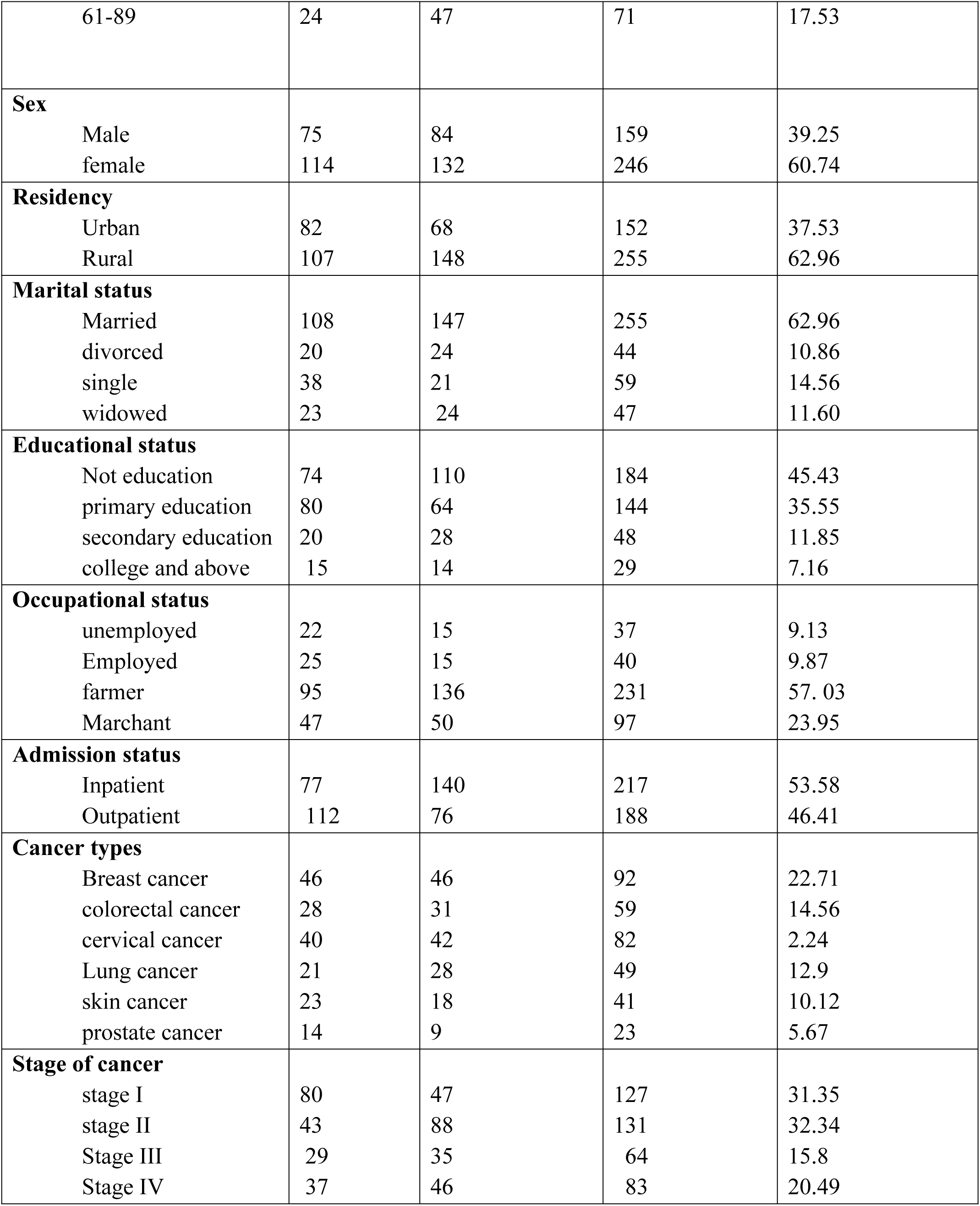
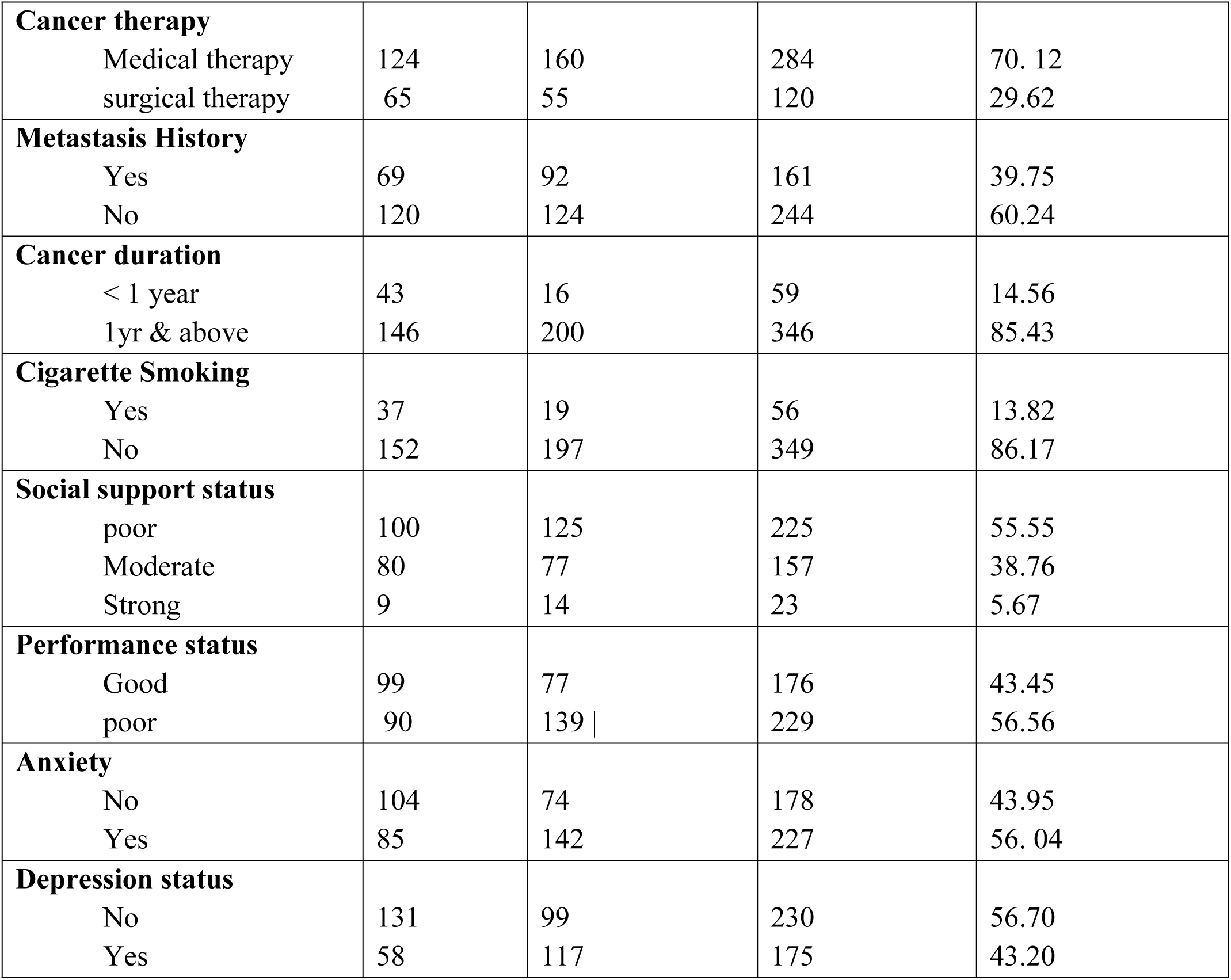
Frequency distribution of the characteristics of study participants attending oncology units in Amhara region comprehensive socialized hospitals, northwest Ethiopia. 2022 (n=405)

| Variable | Fatigue & poor sleep quality<br>comorbidity |  | Frequency | percent (%) |
| --- | --- | --- | --- | --- |
| Age | Yes | No |  |  |
| 18-30 | 42 | 31 | 73 | 18.2 |
| 31-43 | 50 | 45 | 95 | 23.45 |
| 44-60 | 73 | 93 | 166 | 4.98 |
| 61-89 | 24 | 47 | 71 | 17.53 |
| **Sex** |  |  |  |  |
| Male | 75 | 84 | 159 | 39.25 |
| female | 114 | 132 | 246 | 60.74 |
| **Residency** |  |  |  |  |
| Urban | 82 | 68 | 152 | 37.53 |
| Rural | 107 | 148 | 255 | 62.96 |
| **Marital status** |  |  |  |  |
| Married | 108 | 147 | 255 | 62.96 |
| divorced | 20 | 24 | 44 | 10.86 |
| single | 38 | 21 | 59 | 14.56 |
| widowed | 23 | 24 | 47 | 11.60 |
| **Educational status** |  |  |  |  |
| Not education | 74 | 110 | 184 | 45.43 |
| primary education | 80 | 64 | 144 | 35.55 |
| secondary education | 20 | 28 | 48 | 11.85 |
| college and above | 15 | 14 | 29 | 7.16 |
| **Occupational status** |  |  |  |  |
| unemployed | 22 | 15 | 37 | 9.13 |
| Employed | 25 | 15 | 40 | 9.87 |
| farmer | 95 | 136 | 231 | 57.03 |
| Marchant | 47 | 50 | 97 | 23.95 |
| **Admission status** |  |  |  |  |
| Inpatient | 77 | 140 | 217 | 53.58 |
| Outpatient | 112 | 76 | 188 | 46.41 |
| **Cancer types** |  |  |  |  |
| Breast cancer | 46 | 46 | 92 | 22.71 |
| colorectal cancer | 28 | 31 | 59 | 14.56 |
| cervical cancer | 40 | 42 | 82 | 2.24 |
| Lung cancer | 21 | 28 | 49 | 12.9 |
| skin cancer | 23 | 18 | 41 | 10.12 |
| prostate cancer | 14 | 9 | 23 | 5.67 |
| **Stage of cancer** |  |  |  |  |
| stage I | 80 | 47 | 127 | 31.35 |
| stage II | 43 | 88 | 131 | 32.34 |
| Stage III | 29 | 35 | 64 | 15.8 |
| Stage IV | 37 | 46 | 83 | 20.49 |
| **Cancer therapy** |  |  |  |  |
| Medical therapy | 124 | 160 | 284 | 70.12 |
| surgical therapy | 65 | 55 | 120 | 29.62 |
| **Metastasis History** |  |  |  |  |
| Yes | 69 | 92 | 161 | 39.75 |
| No | 120 | 124 | 244 | 60.24 |
| **Cancer duration** |  |  |  |  |
| < 1 year | 43 | 16 | 59 | 14.56 |
| 1yr & above | 146 | 200 | 346 | 85.43 |
| **Cigarette Smoking** |  |  |  |  |
| Yes | 37 | 19 | 56 | 13.82 |
| No | 152 | 197 | 349 | 86.17 |
| **Social support status** |  |  |  |  |
| poor | 100 | 125 | 225 | 55.55 |
| Moderate | 80 | 77 | 157 | 38.76 |
| Strong | 9 | 14 | 23 | 5.67 |
| **Performance status** |  |  |  |  |
| Good | 99 | 77 | 176 | 43.45 |
| poor | 90 | 139 | 229 | 56.56 |
| **Anxiety** |  |  |  |  |
| No | 104 | 74 | 178 | 43.95 |
| Yes | 85 | 142 | 227 | 56.04 |
| **Depression status** |  |  |  |  |
| No | 131 | 99 | 230 | 56.70 |
| Yes | 58 | 117 | 175 | 43.20 |

Magnitudes of comorbidity fatigue and poor sleep quality among adult cancer patient were 46.67% [95% CI: (41.8–51.6] (**figure 2**).

**Fig 2:**
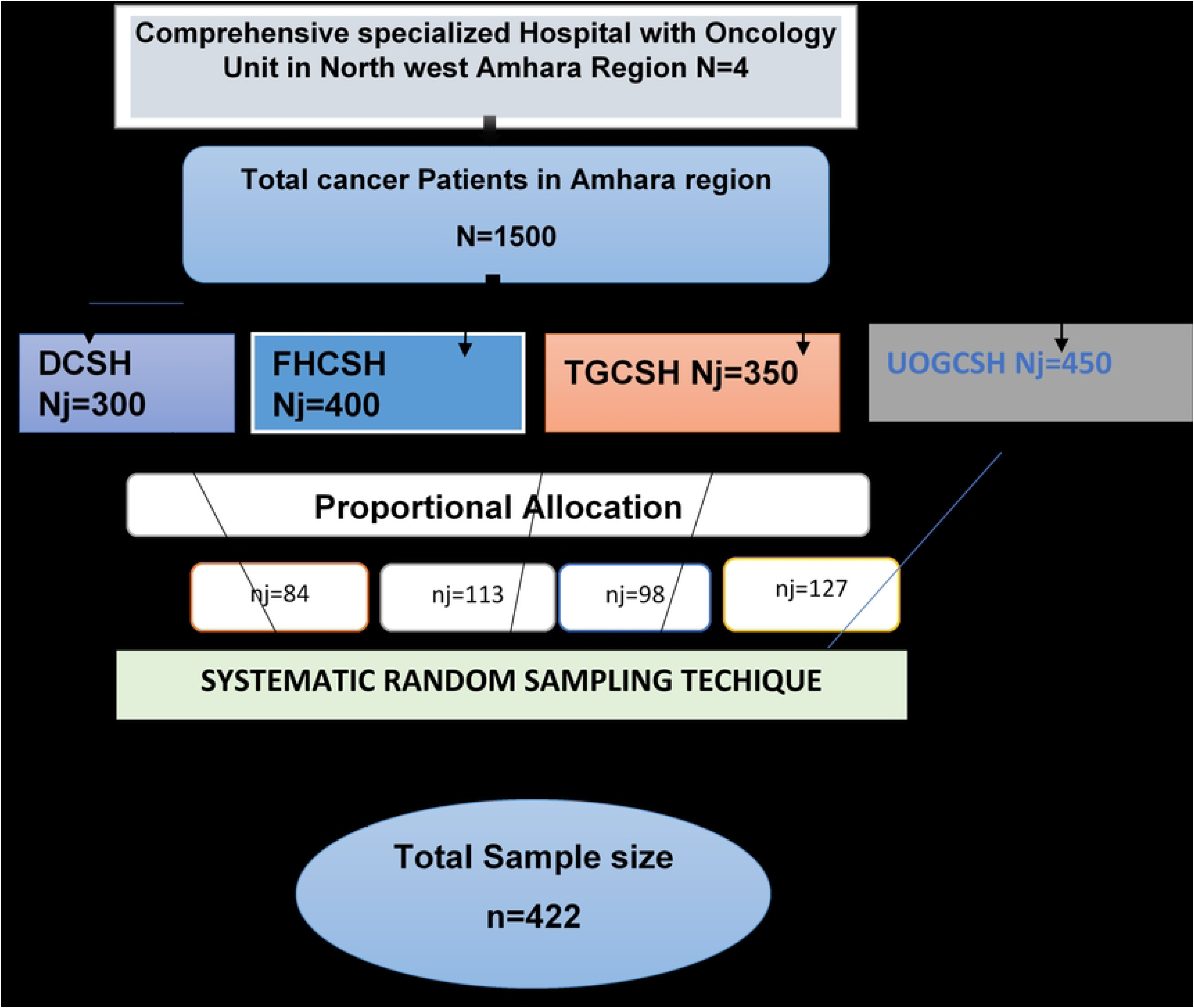
Magnitude of Comorbidity fatigue and poor sleep quality and its predictors among cancer patient in Amhara region, northwest Ethiopia, 2025

### Association factors with comorbidity of fatigue and poor sleep quality among cancer patient Amhara region, northwest, Ethiopia

In the final multivariable logistic regression model, age, marital status, place of residence, admission status, duration of cancer, cancer type, cancer stage, as well as anxiety and depression status were all identified as factors significantly associated with the outcome. The odds of fatigue and poor sleep quality comorbidity were 2.79 times higher among cancer patient aged 61 to 89 years compared to women aged 18 to 30 years [AOR = 2.79, 95% CI: [1.02, 7.62]. The odds of fatigue and poor sleep quality comorbidity were 2.03 times higher among cancer patient living in rural residency compared to urban resident [AOR = 2.03 95%, CI: [1.02, 4.01]. The odds of fatigue and poor sleep quality comorbidity were 2.65 and 3.54 times higher among cancer patient who are married and divorced respectively compared to single cancer patient [AOR= 2.65 95%, CI: (1.01, 6.90)] and [AOR= 3.54 95% CI: (1.10, 11.40)]. The odds of fatigue and poor sleep quality comorbidity were 2.84 times higher among cancer patient who are inpatient compared to outpatient [AOR=2.84, 95%, CI: (1.63, 4.95)]. The odds of fatigue and poor sleep quality comorbidity were 3.92 & 2.52 times higher among Stage II and Stage IV of cancer patient compared to stage I [AOR=3.92, 95%, CI: ([1.89, 8.12] and [AOR= 2.52, 95% CI: (1.04, 6.15)] respectively. The odds of fatigue and poor sleep quality comorbidity were 2.70 times higher among ≥ 1-year cancer duration compared to < l year cancer duration [AOR=2.70, 95% CI: (1.14, 6.39)]. The odds of fatigue and poor sleep quality comorbidity were 1.93 times higher among cancer patient who had an anxiety compared to No anxiety [AOR= 1.93, 95% CI: (1.06, 3.51)]. The odds of fatigue and poor sleep quality comorbidity were 2.10 times higher among cancer patient who had depression compared to No depression [AOR= 2.10, 95% CI: (1.19, 3.70)] (**table 2**).

**Table 2:** Final bivariable and multivariable logistic regression analysis of factors associated with the comorbidity of fatigue and poor sleep quality among cancer patients, Amhara region, northwest Ethiopia.

| Variable | Fatigue & poor sleep quality comorbidity |  | COR [95% CI] | AOR [95% CI] |
| --- | --- | --- | --- | --- |
|  | Yes | No |  |  |
| **Age** |  |  |  |  |
| 18-30 | 42 | 31 | 1 | 1 |
| 31-43 | 50 | 45 | 1.21 [0.65,2.25] | 0.88 [0.38, 2.06] |
| 44-60 | 73 | 93 | 1.72 [0.98, 3.01] | 0.99 [0.44, 2.24] |
| 61-89 | 24 | 47 | 2.65 [1.34, 5.21] | **2.79 [1.02, 7.62] \*** |
| **Sex** |  |  |  |  |
| Female | 75 | 84 | 1 | 1 |
| Male | 114 | 132 | 0.96 [0.64, 1.44] | 0.71 [0.37, 1.37] |
| **Residency** |  |  |  |  |
| Urban | 82 | 68 | 1 |  |
| Rural | 107 | 148 | 1.66 [1.11, 2.50] | **2.03 [1.02, 4.01] \*** |
| **Marital status** |  |  |  |  |
| Single | 108 | 147 | 1 | 1 |
| Married | 20 | 24 | 2.46 [1.36, 4.43] | **2.65 [1.01, 6.90] \*** |
| divorced | 38 | 21 | 2.17 [0.97, 4.82] | **3.54 [1.10, 11.40] \*** |
| widowed | 23 | 24 | 1.88 [.86, 4.12] | 1.12 [0.35, 3.61] |
| **Educational status** |  |  |  |  |
| No education | 74 | 110 | 1 | 1 |
| primary education | 80 | 64 | 0.538 [0.34, 0.83] | 0.85 [0.43, 1.71] |
| secondary edu | 20 | 28 | 0.94 [0.49, 1.79] | 1.88 [0.70, 5.00] |
| college and above | 15 | 14 | 0.62 [0.28, 1.37] | 1.81 [0.55, 5.99] |
| **Occupational status** |  |  |  |  |
| Employed | 22 | 15 | 1 | 1 |
| Unemployed | 25 | 15 | 1.13 [0.45, 2.84] | 0.74 [0.19, 2.94] |
| Farmer | 95 | 136 | 2.38 [1.19, 4.76] | 1.27 [0.43, 3.76] |
| Marchant | 47 | 50 | 1.77 [ 0.83, 3.76] | 1.95 [0.64, 5.99] |
| **Admission status** |  |  |  |  |
| Outpatient | 77 | 140 | 1 |  |
| Inpatient | 112 | 76 | 2.67 [1.79, 4.00] | **2.84 [1.63, 4.95] \*** |
| **Cancer types** |  |  |  |  |
| Breast cancer | 46 | 46 | 1 | 1 |
| colorectal cancer | 28 | 31 | 0.90 [0.46, 1.73] | 1.80 [0.75, 4.34] |
| cervical cancer | 40 | 42 | 0.94 [0.48, 1.85] | 0.91 [0.43, 1.92] |
| lung cancer | 21 | 28 | 1.20[0.56, 2.58] | **3.15 [1.23, 8.03] \*** |
| skin cancer | 23 | 18 | 0.70 [0.31, 1.57] | 1.24 [0.48, 3.22] |
| prostate cancer | 14 | 9 | 0.58 [0.21, 1.54] | 1.14 [0.33,3.88] |
| **Stage of cancer** |  |  |  | 1 |
| stage 1 | 80 | 47 | 1 | **3.92 [1.89, 8.12] \*** |
| stage 2 | 43 | 88 | 3.48 [2.08, 5.81] | 2.33 [0.91, 5.93] |
| stage 3 | 29 | 35 | 2.05 [1.11,3.78] | **2.52 [1.04, 6.15] \*** |
| stage 4 | 37 | 46 | 2.11 [1.20, 3.71] |  |
| **Cancer therapy** |  |  |  |  |
| Medical therapy | 124 | 160 | 1 | 1 |
| surgical therapy | 65 | 55 | 0.65 [.42, 1.00] | 0.69 [0.39, 1.24] |
| **Metastasis History** |  |  |  |  |
| No | 69 | 92 | 1 | 1 |
| yes | 120 | 124 | 1.29 [ 0.86, 1.92] | 1.05 [0.58, 1.90] |
| **Cancer duration** |  |  |  |  |
| < 1 year | 43 | 16 |  | 1 |
| ≥1 year & above | 146 | 200 | 3.68 [1.99, 6.79] | **2.70 [1.14, 6.39] \*** |
| **Cigarette Smoking** |  |  |  |  |
| Yes | 37 | 19 |  | 1 |
| No | 152 | 197 | 2.52 [1.39, 4.56] | 2.06 [0.88, 4.83] |
| **Social support** |  |  |  |  |
| poor | 100 | 125 |  |  |
| moderate | 80 | 77 | 0.77 [0.50, 1.12] | 0.55 [0.31, 0.98] |
| strong | 9 | 14 | 1.24 [0.19, 7.13] | 1.45 [0.12, 16.88] |
| **performance status** |  |  |  |  |
| Good | 99 | 77 | 1 |  |
| poor | 90 | 139 \| | 1.98 [1.33, 2.95] | 0.81 [0.45,1.47] |
| **Anxiety status** |  |  |  |  |
| No | 104 | 74 | 1 |  |
| yes | 85 | 142 | 2.34 [ 1.57, 3.50] | **1.93 [1.06, 3.51] \*** |
| **Depression** |  |  |  |  |
| No | 131 | 99 | 1 |  |
| Yes | 58 | 117 | 2.66 [1.77, 4.01] | **2.10 [1.19, 3.70] \*** |

## Discussion

This study highlights a range of sociodemographic, clinical, and psychological factors associated with the co-occurrence of fatigue and poor sleep quality among individuals with cancer. Findings suggest that certain demographic characteristics, clinical conditions, and psychological states may contribute to the comorbidity of these symptoms. These results offer important insights into the complex interplay of factors that influence symptom burden in cancer populations and provide a foundation for further exploration and targeted interventions.

In this study, comorbidity fatigue and poor sleep quality were 46.67% (95% CI: 41.8–51.6) of adult cancer patients in the Northwest Amhara Region. Our findings are in line with the larger body of research showing that both symptoms are quite common and frequently co-occur in oncology populations, even though no prior studies have specifically recorded this comorbidity as a single outcome. For instance, studies conducted in similar settings have reported that cancer-related fatigue affects approximately 50–60% of patients[16, 33], while poor sleep quality has been observed in 53–61% of patients [16, 34]. The strong associations previously reported between fatigue and sleep problems such as correlation coefficients around 0.6, adjusted odds ratios >2) support the likelihood of a high degree of overlap between the two symptoms[16, 35, 36]. Our finding indicates that around half of cancer patients in this area had both symptoms, which represents a significant symptom burden. Numerous interrelated factors, including the burden of the disease, adverse drug reactions, psychological stress, disease stage, and restricted access to supportive care, may contribute to this. The results highlight the necessity for cancer care teams and doctors to test for sleep disruptions and exhaustion simultaneously, rather than separately, and to think about integrated therapies that treat both symptoms at the same time. However, This magnitude is somewhat lower than reports from Iran 69.3% [37], Arab countries (77.5% and 78%)[15, 38], and Egypt, where fatigue and poor sleep were reported in 99.2% and 87.4% of patients, respectively[39] and America 93% & 77% [40, 41]. Although prior research has repeatedly shown that cancer-related fatigue and poor sleep quality are significantly correlated, most of these studies did not report comorbidity as a single combined prevalence estimate, which makes it difficult to directly compare our findings with those of other studies. Our study’s comparatively lower prevalence could be due to variances in healthcare settings, cultural views of symptoms, study populations, or evaluation instruments. Because fatigue and poor sleep quality can significantly decrease quality of life and may necessitate integrated management techniques in cancer therapy, it is clinically important to recognize their coexistence. The research is still lacking in longitudinal data on the long-term interactions between these two symptoms and if addressing both at the same time with focused therapies can enhance patient outcomes.

In this study, the odds of comorbidity of fatigue and poor sleep quality were 2.79 times higher among cancer patient aged 61 to 89 years compared to women aged 18 to 30 years. This association was consistent with study conducted in of university of California[37, 42], Brazil[43]. Age-related physiological changes including reduced sleep efficiency and more overnight awakenings, higher susceptibility to treatment-related adverse effects, and the existence of numerous comorbidities could all contribute to this outcome. Anxiety, social isolation, and a lack of coping mechanisms are examples of psychosocial issues that can make fatigue and sleep disturbance worse. The combined impact of these variables emphasizes the necessity of focused treatments to enhance sleep and lessen fatigue in elderly cancer patients. And cancer patients living in rural areas had 2.03 times higher odds of comorbid fatigue and poor sleep quality compared to urban residents. This finding is consistent with prior research indicating that rural residency is associated with poorer symptom management, higher fatigue, and greater sleep disturbances among cancer patients[17, 44]. Lower health literacy, longer travel times to clinics, fewer specialized cancer care resources, and restricted access to healthcare services are some potential causes, all of which could lead to a higher cumulative symptom burden and delayed symptom management. Environmental and social variables may make fatigue and sleep issues worse, such as a lack of social support and higher levels of stress in rural areas [45, 46]. The odds of fatigue and poor sleep quality comorbidity were 2.65 and 3.54 times higher among cancer patient who are married and divorced respectively compared to single cancer patient. This finding is consistent with study conducted Rabat, Morocco[47]. Married couples may benefit from psychological and social support, such as emotional support, and encouragement to adhere to treatment, which may have a protective effect on cancer patients’ sleep quality. Support of this kind help in the reduction of stress, worry, and depression symptoms, all of which are closely related to fatigue and sleep quality. On the other hand, cancer patients who are widowed, divorced, or single frequently experience higher levels of psychosocial stress and less coping mechanisms, which raises their risk of fatigue and poor sleep quality[48, 49]. The odds of fatigue and poor sleep quality comorbidity were 2.84 times higher among cancer patient who are inpatient compared to outpatient [AOR=2.84, 95%, CI: (1.63, 4.95)]. This finding was consistent with study conducted in Ethiopia[50], Iran[37], systematic review and meta-analysis[51]. Comorbid fatigue and sleep disturbances are more common in inpatient cancer patients because of the advanced stage of the disease, unmanaged symptoms, and more intense therapy. Sleep is further disrupted and weariness is increased by pain, dyspnea, nausea, and hospital procedures including noise and midnight monitoring. In addition, polypharmacy and psychological discomfort[34, 37]. The odds of fatigue and poor sleep quality comorbidity were 3.92 & 2.52 times higher among Stage II and Stage IV of cancer patient compared to stage I [AOR=3.92, 95%, CI: ([1.89, 8.12] and [AOR= 2.52, 95% CI: (1.04, 6.15)] respectively. This finding was concurrent in study conducted in Ethiopia[10, 16], systematic review and meta-analysis[17]. The higher disease load and physiological stress in patients with more advanced cancer stages increases the likelihood of fatigue and restless nights. Tumor growth in stage II frequently results in elevated inflammatory activity, which throws off circadian rhythm and energy management, making fatigue and sleep quality worse. In stage IV, metastases and widespread disease increase the burden of symptoms, the necessity for intense therapy, and psychological distress, all of which independently affect sleep and cause fatigue. Extensive reviews consistently demonstrate that fatigue and poor sleep quality are strongly predicted by tumor stage, symptom burden, and treatment severity [34, 52]. The odds of fatigue and poor sleep quality comorbidity were 2.70 times higher among ≥ 1-year cancer duration compared to < l year cancer duration [AOR=2.70, 95% CI: (1.14, 6.39)]. This finding was concurrent with study conducted in USA[53], Netherlands[54], Ethiopia[16]. Patients who have had cancer for more than a year are far more likely than those who have had it for less time to suffer from concomitant fatigue and poor sleep quality. Long-term exposure to the disease and its treatments might result in cumulative adverse effects from radiation, chemotherapy, or surgery, which can worsen sleep problems and exhaustion. Furthermore, extended cancer duration is frequently linked to higher levels of psychological distress, such as anxiety and depression, as well as a greater load of symptoms, such as pain and discomfort, all of which worsen energy and sleep quality. The odds of fatigue and poor sleep quality comorbidity were 1.93 times higher among cancer patient who had an anxiety compared to No anxiety [AOR= 1.93, 95% CI: (1.06, 3.51)]. This associate was similar with study done in China[34], Ethiopia[50], meta-analysis studies in multiple countries[51]. Cancer patients’ comorbidity of fatigue and poor sleep quality is greatly influenced by anxiety through both physiological and psychological factors. It causes the hypothalamic-pituitary-adrenal (HPA) axis to become dysregulated and hyperarousal, which interferes with sleep and results in non-restorative slumber. Consequently, this makes cancer-related fatigue worse, which is a prevalent and enduring symptom in oncology populations. Additionally, anxiety exacerbates fatigue and sleep disruptions by reducing adaptive coping mechanisms [55–57].

The odds of fatigue and poor sleep quality comorbidity were 2.10 times higher among cancer patient who had depression compared to No depression [AOR= 2.10, 95% CI: (1.19, 3.70)]. This finding concurrent with study conducted in Ethiopia and Egypt [9, 39, 58].

There are several interrelated biological and psychological processes via which depression in cancer patients can worsen fatigue and interfere with sleep. Increased exhaustion results from neurochemical imbalances linked to depression, such as changed serotonin and dopamine levels, which interfere with sleep regulation and decrease restorative sleep. Moreover, depression frequently entails dysregulation of the hypothalamic-pituitary-adrenal (HPA) axis and increased inflammatory responses, which exacerbate fatigue and poor sleep quality. On a psychological level, depression symptoms make people less motivated and energetic, which makes it harder for them to take care of themselves and keep up good sleep habits. This makes the symptoms worse. All of these processes work together to explain why depressed cancer patients have been shown to have higher risks of experiencing fatigue and poor sleep comorbidities [39, 59].

## Strengths and Limitations

The major strengths of this study its novel focus on comorbidity, addressing a research gap by assessing the combined prevalence of fatigue and poor sleep in a resource-limited setting. Additionally, the use of validated assessment tools for both fatigue and sleep quality enhances the reliability of our findings. The study’s relatively large and diverse sample drawn from multiple facilities in the Northwest Amhara Region improves its representativeness and relevance to clinical practice. However, several limitations should be acknowledged; the reliance on self-reported data may introduce recall bias, particularly in the subjective evaluation of symptoms, the absence of biochemical or clinical staging data limits the ability to assess the influence of disease severity or treatment modality on symptom burden, while we used a binary outcome to capture comorbidity, more nuanced analyses such as symptom cluster modeling or longitudinal tracking may yield deeper insights into symptom interactions over time.

## Clinical Implications

Nearly half of adult cancer patients in the region experience both fatigue and poor sleep quality, highlighting the need for routine screening of concurrent symptoms. These comorbidities intensify each other’s effects, leading to impaired functioning, reduced quality of life, and lower treatment adherence. Integrated management strategies ranging from CBT and physical activity programs to basic sleep hygiene counseling are essential, especially in resource-limited settings.

## Conclusions and Recommendation

Comorbid fatigue and poor sleep quality are highly prevalent among adult cancer patients in the Northwest Amhara Region, significantly affecting functional status and quality of life. These findings underscore the need for routine screening and integrated management of concurrent symptoms within oncology care. Simple interventions such as sleep hygiene counseling, psychoeducation, and targeted therapies can improve patient outcomes, especially in resource-limited settings. Future research should explore longitudinal trajectories and contributing factors to inform tailored, effective interventions.

## Declaration

### Clinical trial number

Not applicable

### Ethical consideration

This study was conducted in accordance with the ethical principles of the Declaration of Helsinki. Ethical approval was obtained from the Research and Ethical Review Committee of the University of Gondar, College of Medicine and Health Sciences, School of Nursing (Reference No. S/N 237/2014). The committee approved the study on behalf of the Institutional Ethical Review Committee of the University of Gondar. The objectives and significance of the study were clearly explained to all participants, and written informed consent was obtained from each participant prior to data collection.

## Author’s contributions

**GAZ**: involved data collection, data analysis, interpretation, report and manuscript writing.

**AMB:** involved in analysis, and result interpretation

**BAN:** involved in data analysis, interpretation, and, and manuscript writing.

**DGA:** involved in discussion and drafting proposal and interpretation of result.

**YGB:** involved in conceptualization, validation, writing original draft.

**AEB:** involved in data collection and writing original draft

**AFZ:** involved in designing and preparing manuscript.

## Data Availability

All relevant data are within the manuscript and its Supporting Information files

The manuscript contains all of the data that is crucial to our findings. Request for additional information on the data set and questions about data sharing will be treated in accordance with a reasonable request to.

